# Genomic and Virological Characterization of SARS-CoV-2 Variants in a Subset of Unvaccinated and Vaccinated U.S. Military Personnel

**DOI:** 10.1101/2021.12.16.21267862

**Authors:** Darci R. Smith, Christopher Singh, Jennetta Green, Matthew R. Lueder, Catherine E. Arnold, Logan J. Voegtly, Kyle A. Long, Gregory K. Rice, Andrea Luquette, Haven Miner, Lindsay Glang, Andrew Bennett, Robin Miller, Francisco Malagon, Regina Z. Cer, Kimberly A. Bishop-Lilly

## Abstract

The emergence of SARS-CoV-2 variants complicates efforts to control the COVID-19 pandemic. Increasing genomic surveillance of SARS-CoV-2 is imperative for early detection of emerging variants, to trace the movement of variants, and to monitor effectiveness of countermeasures. Additionally, determining the amount of viable virus present in clinical samples is helpful to better understand the impact these variants have on viral shedding. In this study, we analyzed nasal swab samples collected between March 2020 and early November 2021 from a cohort of United States (U.S.) military personnel and healthcare system beneficiaries stationed worldwide as a part of the Defense Health Agency’s (DHA) Global Emerging Infections Surveillance (GEIS) program. SARS-CoV-2 quantitative real time reverse-transcription PCR (qRT-PCR) positive samples were characterized by next-generation sequencing and a subset was analyzed for isolation and quantification of viable virus. Not surprisingly, we found that the Delta variant is the predominant strain circulating among U.S. military personnel beginning in July 2021 and primarily represents cases of vaccine breakthrough infections (VBIs). Among VBIs, we found a 50-fold increase in viable virus in nasal swab samples from Delta variant cases when compared to cases involving other variants. Notably, we found a 40-fold increase in viable virus in nasal swab samples from VBIs involving Delta as compared to unvaccinated personnel infected with other variants prior to the availability of approved vaccines. This study provides important insight about the genomic and virological characterization of SARS-CoV-2 isolates from a unique study population with a global presence.

**Impact:** The COVID-19 pandemic is currently a leading cause of death globally and new SARS-CoV-2 variants continue to emerge. Genomic surveillance of variants is necessary to characterize mutations that could affect transmissibility and spread, antigenicity or virulence. Furthermore, wet lab studies are necessary to evaluate how genetic differences may affect viral fitness and transmissibility. The Delta variant is currently the predominant strain globally and determining factors that drive its ability to spread rapidly is important. In this study, we characterized SARS-CoV-2 positive samples from U.S. military personnel and their beneficiaries by next-generation sequencing and isolation of viable virus. Consistent with other studies, we found higher levels of infectious virus from Delta samples when compared to non-Delta infections. Strikingly, we found the difference in titer between Delta and other strains to be so profound as to be unaffected by vaccination status, suggesting that increased transmissibility of the Delta variant is in part due to higher amounts of virus shedding. This helps explain the rapid spread of the Delta variant and provides the impetus to increase control measures such as vaccination, boosters, masking and distancing requirements. It will be necessary to continue genomic and virological characterization of new variants, such as Omicron.

## Introduction

The emergence of severe acute respiratory syndrome coronavirus 2 (SARS-CoV-2), the causative agent of coronavirus disease 2019 (COVID-19), has led to a historic global pandemic. Multiple SARS-CoV-2 genetic variants are circulating globally, some of which have mutations of concern or mutations of interest that could impact transmissibility and spread, antigenicity or virulence (1). The World Health Organization (WHO) classifies many of these as variants of concern (VOC) or variants of interest (VOI) based on mutations that may impact SARS-CoV-2 countermeasures, including vaccines, therapeutics, and diagnostics. Current VOC as defined by the WHO include B.1.1.7 (Alpha), B.1.351 (Beta), P.1 (Gamma), B.1.617.2 (Delta), and most recently, B.1.1.529 (Omicron), which was designated as a VOC on 26 November 2021. The Alpha variant was initially detected in the United Kingdom at the end of 2020 and remained as the dominant strain globally until May 2021 (2). The Delta variant was first detected in India and became a VOI in April 2021 and a VOC in May 2021 when it became the dominant strain identified worldwide (3). The Delta variant is estimated to be more transmissible than the Alpha variant, is less sensitive to neutralizing antibodies, and causes increased hospitalization rates mainly in unvaccinated individuals (4-6). There are currently multiple vaccines in use either with FDA approval or emergency use authorization (EUA) that have shown efficacy, however vaccine breakthrough infections (VBIs) do occur and may increase with the emergence of novel variants (7).

Quantitative real time reverse-transcription PCR (qRT-PCR) is the benchmark method for COVID-19 diagnosis, but an important limitation of this approach is the assay only detects RNA and does not detect infectious virus. This can be problematic since several studies have described the persistence of SARS-CoV-2 RNA within different body sites (8-10) and viral RNA levels as a measurement of viral load does not always correlate to the amount of viable virus in a given sample. Quantitative assays that measure replication competent virus are important for determining infectious viral titers in clinical samples, which is critical to informing infection prevention and control guidelines for current and future SARS-CoV-2 variants.

Detection of infectious virus is determined by *in vitro* viral culture on susceptible cell lines. However, the ability of viral culture methods to provide insight on infectious shedding is hampered by the fact that this method is labor-intensive and requires biosafety level 3 (BSL-3) facilities for SARS-CoV-2. There are also assay variables such as the cell line and methods used to quantify the amount of virus. Vero cells and their subclones or derivatives are commonly used to culture SARS-CoV-2 and Vero E6 cells expressing human transmembrane serine protease 2 (TMPRSS2) have demonstrated enhanced susceptibility and isolation of SARS-CoV-2 (11). Plaque or median tissue culture infectious dose (TCID_50_) assays are commonly used to quantify viable virus, but new methods have been described like the S-Fuse assay that uses a cell reporter system and is based on the detection of GFP positive syncytia formation between infected and neighboring cells (12).

While some studies have described the amount of viable virus shed by individuals infected with the various SARS-CoV-2 variants (12-14), more studies are needed to better characterize infectious virus levels in vaccinated and unvaccinated individuals. This is especially important because each study uses different methods and focuses on different populations of individuals from different geographic locations. In this study, we analyzed nasal swab samples collected between March 2020 and early November 2021 from a cohort of U.S. military personnel and beneficiaries stationed worldwide as a part of the DHA’s GEIS program. SARS-CoV-2 qRT-PCR positive samples were characterized by next-generation sequencing and a subset was analyzed for isolation and quantification of viable virus. Following the EUA approval of the COVID-19 vaccines, samples were collected from vaccinated individuals to characterize VBIs. This study provides unique insight about the genomic and virological characteristics of SARS-CoV-2 isolates from a generally healthy population with a global presence that spans from the beginning of the pandemic to early November 2021.

## Methods

### Genome sequencing

Viral genome sequencing was conducted on the Illumina MiSeq platform using the ARTIC nCoV-2019 Sequencing protocol (15) and the YouSeq SARS-CoV-2 Coronavirus NGS Library prep kit (YouSeq). Approximately 100ng of RNA was reverse-transcribed as in the protocol, however the YouSeq reverse transcriptase was replaced with SuperScript IV (ThermoFisher Scientific). cDNA was amplified using multiplex PCR and either the associated ARTIC primer pools or YouSeq primer pools. Libraries prepared via the ARTIC protocol were cleaned using 1x AMPure XP beads (Beckman Coulter) and re-suspended in nuclease free molecular grade water. The samples were then processed following the QiaSeq FX protocol (Qiagen) and libraries were completed. Libraries were quality-checked using an Agilent Bioanalyzer High sensitivity kit (Agilent) and quantified using the Qubit DNA High Sensitivity assay (ThermoFisher Scientific) prior to sequencing using Illumina MiSeq v3 2×300 chemistry (Illumina).

### Bioinformatics Analyses

Viral Amplicon Illumina Workflow (VAIW) was used to collate SARS-CoV-2 consensus genomes (16). Briefly, Illumina reads were quality trimmed and filtered to >Q20 and minimum length of 50 bp using bbmap (17). Paired reads were merged using bbmerge with default settings (18). Trimmed, filtered, and merged reads were aligned to the Wuhan reference genome (NCBI GenBank accession NC_045512.2/MN908947.3) using bbmap with local alignment and maximum insertion/deletion of 500 bp (17). Primers were trimmed from sequences using align_trim from ARTIC workflow/pipeline (19). Once a high quality consensus genome was obtained, Single Nucleotide Variants (SNVs) were determined using SAMtools mpileup (20) and iVar (Intrahost variant analysis of replicates) (21) using a minimum frequency of 0.3 and a minimum read depth of 10 (22).(23-25) Lineage information was derived using Pangolin (Phylogenetic Assignment of Named Global Outbreak LINeages) v3.1.16; github.com/hCoV-2019/pangolin) (26)

### Virus culture

Patient samples were cultured for SARS-CoV-2 using a standard plaque assay on Vero E6 cells expressing TMPRSS2 [4] in six-well plates and a cytopathic effect (CPE) assay on Vero E6/TMPRSS2 cells in T25 cm^2^ flasks. For the plaque assay, duplicate wells were infected with 0.2mL aliquots from serial 10-fold dilutions in Minimum Essential Medium (MEM) followed by a 1 hr incubation at 37°C, 5% CO_2_ to allow virus adsorption to occur. After incubation, cells were overlaid with MEM containing 0.5% agar supplemented with 5% heat-inactivated FBS, 1% Penicillin/Streptomycin and incubated for 48 hr at 37 °C, 5% CO_2_. Cells were fixed in 10% formalin prior to staining with crystal violet. For the CPE assay, cells were seeded in T25 cm^2^ flasks and each flask was infected with 0.5mL aliquots from a 1:10 dilution in Dulbecco’s Modified Eagle Medium (DMEM) followed by a 1 hr incubation at 37°C, 5% CO_2_. After incubation, 5mL of DMEM supplemented with 5% heat-inactivated FBS, 1% Penicillin/Streptomycin was incubated for 4 days at 37°C, 5% CO_2_. CPE was monitored daily. After 4 days, supernatant was passed onto fresh cells to allow additional time to amplify. The presence of SARS-CoV-2 was confirmed by lateral flow immunoassay using the Quidel Sofia 2 analyzer and/or qRT-PCR.

### Statistical analysis

Statistical analysis was performed using GraphPad Prism. Tukey’s multiple comparisons test was used to compare the amount of viable virus between the unvaccinated and vaccinated groups.

## Results

We have sequenced over 2,300 nasal swab samples collected from March 2020 until early November 2021 and of those, we produced 1,304 coding complete genomes for which we then determined the Pango lineages (Figure 1). Smaller numbers of samples were received early in the pandemic compared to after the variants began to emerge when there was more impetus to sequence a greater number of samples. The first samples sequenced in March and April 2020 were similar to strains at the root of the pandemic (lineage A and sublineages) and are classified as A.1, A.3 etc., collectively designated as Pango lineage “other” in Figure 1A and C. The “other” category also contains B lineages and was used for any lineage with 12 or less representatives. The large European lineage B.1 was detected in six samples in April 2020. In February 2021, there was a strong commitment within the DoD to sequence a greater number of samples with the rationale that it is important to continually perform genomic surveillance of the SARS-CoV-2 genome for early detection of emerging variants and to monitor variant cases in several geographic locations. The VOC B.1.1.7 (Alpha) was the predominant strain detected between February and June 2021. Many military personnel were considered fully vaccinated during this same timeframe and some of these samples represent cases of VBI. The former VOC, B.1.526 (Iota), which is a lineage predominantly circulating in New York, was detected from March to June 2021 in 29 samples. The VOC, B.1.617.2 (Delta) was first detected in May 2021 in one sample collected from Portsmouth, Virginia. An increasing number of B.1.617.2 (Delta) was detected starting in June 2021 and by July 2021 was the dominant variant detected in all samples received. Multiple sublineages of B.1.617.2 were detected which increased over time (Figure 1B and D). The “other” category for Figure 1B and D was used for any sublineage with 5 or less representatives. The Omicron variant was not detected during the timeframe of our current study.

**Figure 1:**
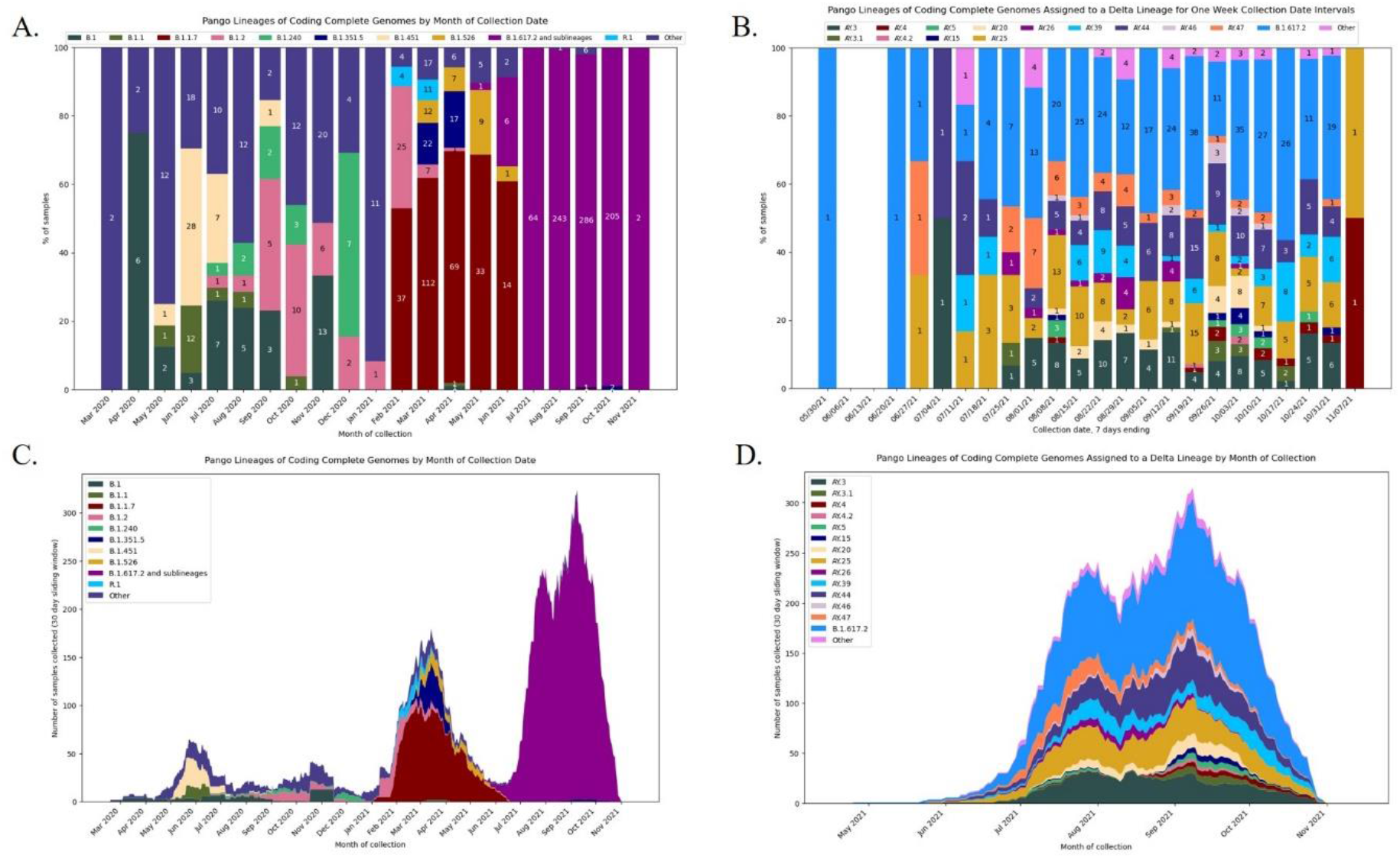
Pango lineages of circulating SARS-CoV-2 strains in the U.S. military before and after implementation of the EUA vaccines. A) Distribution of Pango lineages by month for all samples; B) distribution of Delta variant sublineages by month; C) stacked chart displaying all lineages per month; D) stacked chart displaying Delta variant sublineages by month.

A subset of samples were selected to determine if viable virus was present where we compared viral titers in samples collected from unvaccinated individuals and cases of VBI (Figure 2). A summary of the results broken down by Pango lineage is shown in Table 1 and 2 for unvaccinated and vaccinated individuals, respectively. We detected viable virus in 25 samples received from unvaccinated personnel collected between March 2020 and January 2021, which is prior to the emergence of VOCs. The mean amount of virus detected in unvaccinated personnel was 3.2 log_10_ PFU/mL, which is similar to the amount of virus detected from VBIs associated with all variants except for Delta (3.1 log_10_ PFU/mL). Significantly more virus was detected in VBIs associated with the Delta variant when compared to unvaccinated personnel.

**Figure 2:**
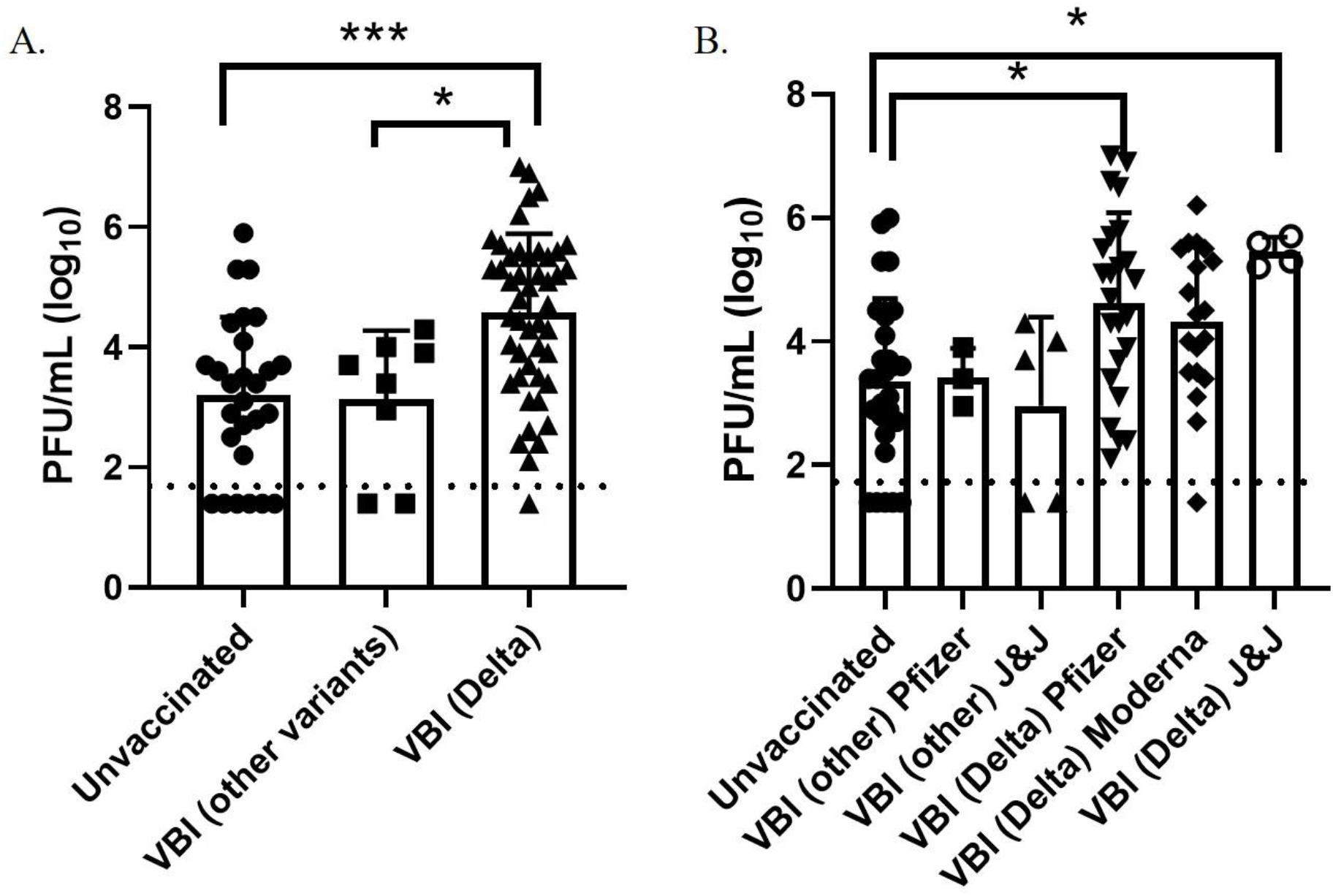
Infectious virus titers in unvaccinated and vaccinated personnel. A) Viral titers in nasal swabs determined by standard plaque assay; B) viral titers broken down by vaccine type. Error bars represent the standard deviation, and the dashed line represents the detection limit of the plaque assay. Symbols below this line were negative by the plaque assay, but positive by the CPE assay. Asterisks indicate statistical significance determined by Tukey’s multiple comparisons test.

**Table 1:** Summary of viable virus detection in nasal swab samples from unvaccinated individuals broken down by Pango lineage.

| Pango Lineage | Number of Samples | Range of Viable Virus (mean); PFU/mL (log<sub>10</sub>) |
| --- | --- | --- |
| **B.1.2** | 1 | 3.0 |
| **B.1** | 18 | 1.4-5.9 (4.9) |
| **B.1.1.9** | 4 | 1.4-3.7 (3.3) |
| **B.6** | 2 | 2.8-2.9 (2.85) |

**Table 2:**
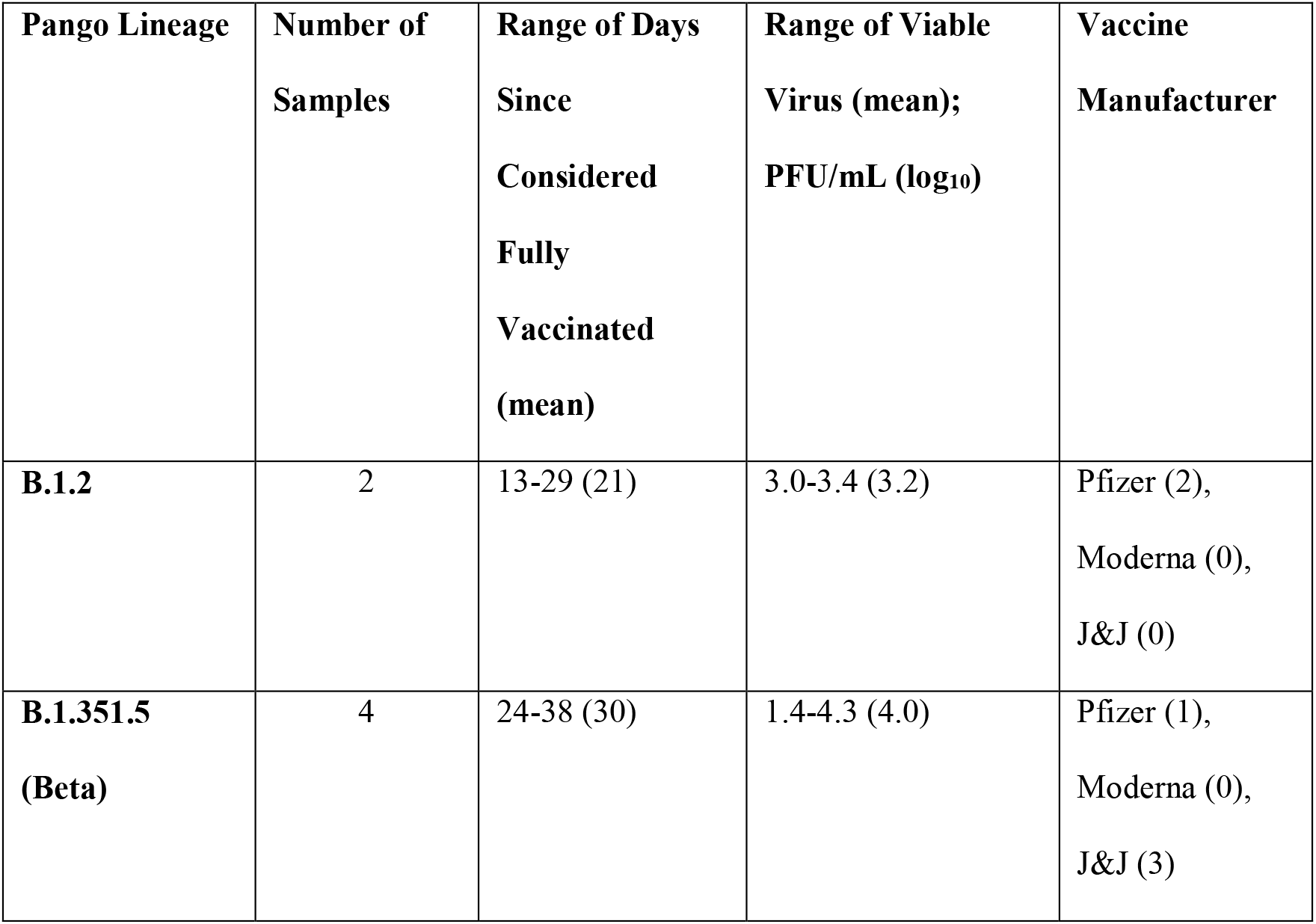
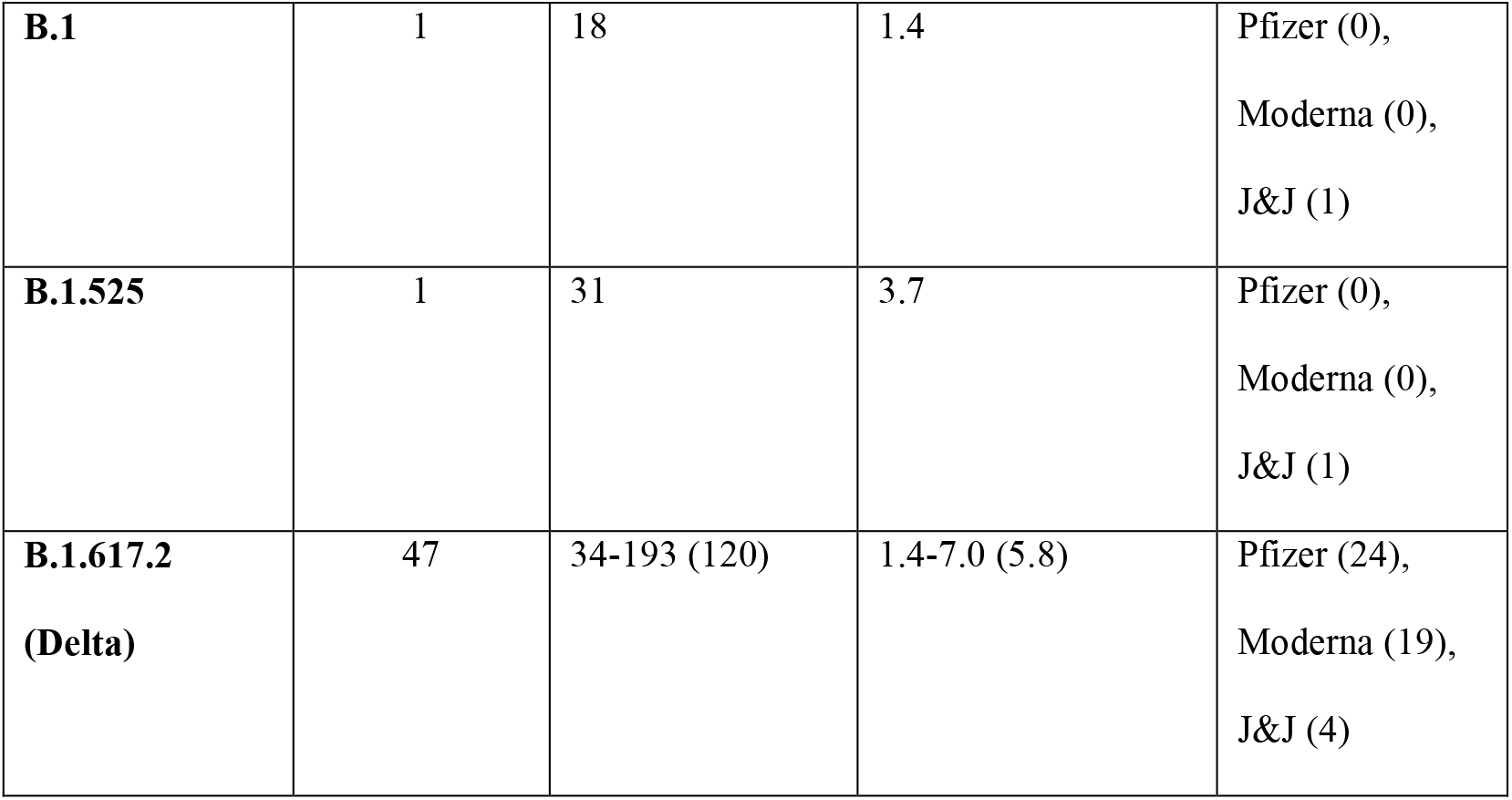
Summary of viable virus detection in nasal swab samples from vaccinated individuals broken down by Pango lineage.

The mean amount of virus detected in VBIs associated with the Delta variant was 4.6 log_10_ PFU/mL, which was statistically significant by Tukey’s multiple comparisons test (p=0.0001) when compared to the amount of virus detected in unvaccinated personnel prior to the emergence of VOCs. In contrast to the unvaccinated samples, almost all the VBI samples associated with the Delta variant had detectable virus that was above the level of detection for the plaque assay and one sample had a titer as high as 7 log_10_ PFU/mL. Samples from unvaccinated personnel infected with the Delta variant were not available for this study since the majority of military personnel were fully vaccinated when the Delta variant became the predominant strain. The amount of virus detected in VBIs associated with the Delta variant compared to VBIs with other variants was also statistically significant by Tukey’s multiple comparisons test (p=0.0132) although with less power because of the low number of VBI samples associated with other variants. The results are further broken down by vaccine manufacturer (Figure 2B) where we continued to observe a statistically significant difference in viral titers for unvaccinated individuals with non-Delta variants compared to samples from VBI associated with the Delta variant from individuals that received the Pfizer (p=0.0106) and Johnson & Johnson (p=0.0427) vaccines. Interestingly, VBI associated with the Delta variant that received the Moderna vaccine was not statistically significant when compared to the unvaccinated group. This may be a reflection of the limited number of samples in our analyses.

## Discussion

The Delta variant displaced the Alpha variant and was identified as the predominant strain in military personnel starting in July 2021. Currently, multiple sublineages of B.1.617.2 continue to be identified. A greater proportion of the VBI cases in our dataset have been associated with the Delta variant, commensurate with its current preponderance in the population at this point in time. We quantified the amount of viable virus in a subset of samples, and our results suggest that the increased transmissibility of the Delta variant is in part due to higher amounts of virus shedding. We detected 50-fold more infectious virus in VBIs involving Delta compared to VBIs with other variants. We also detected 40-fold more infectious virus in VBIs involving Delta compared to unvaccinated personnel infected with non-Delta variants.

Our results are similar to what has been reported for the rest of the population in the U.S. where the Delta variant is currently the predominant strain and is more than twice as infectious as previous strains. The improved viral fitness of the Delta variant has been associated with mutations in the furin cleavage site that allow the virus to enter cells more efficiently (27, 28). Another study found that four mutations in the nucleocapsid (N) protein, which is an important protein for viral replication, improve the viruses’ ability to form infectious particles. These four mutations in N, which are universally found in more transmissible SARS-CoV-2 variants, increased mRNA delivery and expression and two of the mutations produced more infectious virus (29). This is consistent with other studies that found that samples from people infected with the Delta variant have higher viral RNA levels and an increased duration of viral shedding compared to samples collected from people infected with other variants (14, 30-32). Furthermore, the levels of viral RNA from infected individuals have been shown to be similar for vaccinated and unvaccinated individuals (14, 33).

Numerous studies measure viral load by the amount of viral RNA in a clinical sample, but do not determine the amount of viable virus. This is likely due to the cumbersome nature of the assays used to quantify levels of infectious virus coupled with the requirement to perform these assays in a BSL-3 laboratory. However, a recent study found a significant amount of variability in the relationship between viral titers for infectious virus and viral RNA levels (13). Their results indicate that determining viral load by RNA levels in clinical samples may have limitations and assays for viable virus should be included for emerging variants. Additionally, they detected six times as much infectious virus for the same amount of RNA for Delta variant samples compared to Alpha variant samples. Our results are similar where we found that the Delta variant cases shed viable virus at higher levels. Similar levels of virus were detected from unvaccinated personnel with non-Delta infections prior to the availability of vaccines granted EUA and FDA approval compared to VBI due to other variants besides Delta. In contrast, 50-fold more infectious virus was detected in samples from VBI due to Delta compared to VBIs with other variants. When we analyzed our results by vaccine manufacturer, we continued to detect significantly higher levels of infectious virus in samples from individuals who received the Pfizer and Johnson & Johnson vaccines and experienced a VBI due to Delta when compared to the unvaccinated personnel with non-Delta infections. Interestingly, VBI associated with the Delta variant that received the Moderna vaccine was not statistically significant when compared to the unvaccinated group. This may be a reflection of the limited number of samples in our analyses. Another limitation of our study is the lack of access to metadata for the clinical specimens, which affects our interpretation of the results by differences in age, pre-existing conditions, and days from exposure or symptom onset. However, most of our samples did come from active duty military personnel stationed worldwide who are considered young (age 18-40), are required to maintain a physical fitness regimen and pass routine physicals, and are therefore considered relatively healthy compared to the general population.

Other studies have also found that viable virus is more likely to be isolated from clinical samples from Delta compared to Alpha (14). The authors do point out that there was a significantly longer period between being considered fully vaccinated and when VBI with the Delta variant occurred, which is similar to our results. However, our data demonstrating that we detected 40-fold more infectious virus in VBIs involving Delta compared to unvaccinated personnel with non-Delta infections prior to the availability of vaccines granted EUA and FDA approval suggests that the increase in the amount of viable virus is due to the variant and not the vaccination status. This is consistent with the study by Luo *et al*. that found the recovery of infectious virus was significantly higher with the Delta variant compared to Alpha in both vaccinated and unvaccinated groups (14).

Our results highlight the importance of characterizing the SARS-CoV-2 genome and infectious viral titers of emerging variants isolated from unvaccinated and vaccinated personnel. This work contributes to the growing body of evidence that the increased fitness of Delta is primarily due to viral genetic determinants that increase infectivity or replication. This helps to explain the rapid spread of the Delta variant and provides the impetus to increase control measures such as vaccination, boosters, masking and distancing requirements to prevent ongoing transmission. Furthermore, this study demonstrates the value of pairing genomic and virological characterization when assessing novel SARS-CoV-2 variants. These parallel approaches should be applied to newly emerging variants, such as Omicron, and other variants that may arise in the future.

## Disclaimer

The views expressed in this article are those of the authors and do not necessarily reflect the official policy or position of the Department of Defense, the Navy, or the U.S. Government. Several of the authors are U.S. Government employees. This work was prepared as part of their official duties. Title 17 U.S.C. § 105 provides that ‘Copyright protection under this title is not available for any work of the United States Government.’ Title 17 U.S.C. §101 defines a U.S. Government work as a work prepared by a military service member or employee of the U.S. Government as part of that person’s official duties.

## Data Availability

All data produced in the present work are contained in the manuscript or available upon reasonable request to the authors.

## Acknowledgements

We thank Justice Akuoku Frimpong, Kevin Schully, and Christina Farris for technical assistance with the viral cultures. This study was supported by the Armed Forces Health Surveillance Division (AFHSD), Global Emerging Infections Surveillance (GEIS) Branch, ProMIS IDs P0013_AH_01.01, P0166_20_NM, P0093_21_NM, and P0199_21_NM to KABL as well as Navy Work Unit Number (WUN) A1417.

## References

1. Hadj Hassine I. Covid-19 vaccines and variants of concern: A review. Rev Med Virol (2021):e2313. Epub 2021/11/11. doi: 10.1002/rmv.2313. PubMed PMID: 34755408.

2. Davies NG, Abbott S, Barnard RC, Jarvis CI, Kucharski AJ, Munday JD, et al. Estimated transmissibility and impact of SARS-CoV-2 lineage B.1.1.7 in England. Science (2021) 372(6538). Epub 2021/03/05. doi: 10.1126/science.abg3055. PubMed PMID: 33658326; PubMed Central PMCID: PMCPMC8128288.

3. GISAID. hCov19 Variants [cited 2021 December 1]. Available from: https://www.gisaid.org/hcov19-variants/.

4. Planas D, Veyer D, Baidaliuk A, Staropoli I, Guivel-Benhassine F, Rajah MM, et al. Reduced sensitivity of SARS-CoV-2 variant Delta to antibody neutralization. Nature (2021) 596(7871):276–80. Epub 2021/07/09. doi: 10.1038/s41586-021-03777-9. PubMed PMID: 34237773.

5. Pouwels KB, Pritchard E, Matthews PC, Stoesser N, Eyre DW, Vihta KD, et al. Effect of Delta variant on viral burden and vaccine effectiveness against new SARS-CoV-2 infections in the UK. Nat Med (2021). Epub 2021/10/16. doi: 10.1038/s41591-021-01548-7. PubMed PMID: 34650248.

6. Twohig KA, Nyberg T, Zaidi A, Thelwall S, Sinnathamby MA, Aliabadi S, et al. Hospital admission and emergency care attendance risk for SARS-CoV-2 delta (B.1.617.2) compared with alpha (B.1.1.7) variants of concern: a cohort study. Lancet Infect Dis (2021). Epub 2021/08/31. doi: 10.1016/S1473-3099(21)00475-8. PubMed PMID: 34461056; PubMed Central PMCID: PMCPMC8397301 funding from GlaxoSmithKline for a research project related to seasonal influenza and antiviral treatment; this project preceded and had no relation to COVID-19, and GD had no role in and received no funding from the project. All other authors declare no competing interests.

7. Lipsitch M, Krammer F, Regev-Yochay G, Lustig Y, Balicer RD. SARS-CoV-2 breakthrough infections in vaccinated individuals: measurement, causes and impact. Nature Reviews Immunology (2021). doi: 10.1038/s41577-021-00662-4.

8. He X, Lau EHY, Wu P, Deng X, Wang J, Hao X, et al. Temporal dynamics in viral shedding and transmissibility of COVID-19. Nat Med (2020) 26(5):672–5. Epub 2020/04/17. doi: 10.1038/s41591-020-0869-5. PubMed PMID: 32296168.

9. Wolfel R, Corman VM, Guggemos W, Seilmaier M, Zange S, Muller MA, et al. Virological assessment of hospitalized patients with COVID-2019. Nature (2020) 581(7809):465–9. Epub 2020/04/03. doi: 10.1038/s41586-020-2196-x. PubMed PMID: 32235945.

10. Zapor M. Persistent Detection and Infectious Potential of SARS-CoV-2 Virus in Clinical Specimens from COVID-19 Patients. Viruses (2020) 12(12). Epub 2020/12/09. doi: 10.3390/v12121384. PubMed PMID: 33287245; PubMed Central PMCID: PMCPMC7761721.

11. Matsuyama S, Nao N, Shirato K, Kawase M, Saito S, Takayama I, et al. Enhanced isolation of SARS-CoV-2 by TMPRSS2-expressing cells. Proc Natl Acad Sci U S A (2020) 117(13):7001–3. Epub 2020/03/14. doi: 10.1073/pnas.2002589117. PubMed PMID: 32165541; PubMed Central PMCID: PMCPMC7132130.

12. Monel B, Planas D, Grzelak L, Smith N, Robillard N, Staropoli I, et al. Release of infectious virus and cytokines in nasopharyngeal swabs from individuals infected with non-alpha or alpha SARS-CoV-2 variants: an observational retrospective study. EBioMedicine (2021) 73:103637. Epub 2021/10/23. doi: 10.1016/j.ebiom.2021.103637. PubMed PMID: 34678613; PubMed Central PMCID: PMCPMC8526063.

13. Despres HW, Mills MG, Shirley DJ, Schmidt MM, Huang ML, Jerome KR, et al. Quantitative measurement of infectious virus in SARS-CoV-2 Alpha, Delta and Epsilon variants reveals higher infectivity (viral titer:RNA ratio) in clinical samples containing the Delta and Epsilon variants. medRxiv (2021). Epub 2021/09/29. doi: 10.1101/2021.09.07.21263229. PubMed PMID: 34580674; PubMed Central PMCID: PMCPMC8475961.

14. Luo CH, Morris CP, Sachithanandham J, Amadi A, Gaston D, Li M, et al. Infection with the SARS-CoV-2 Delta Variant is Associated with Higher Infectious Virus Loads Compared to the Alpha Variant in both Unvaccinated and Vaccinated Individuals. medRxiv (2021). Epub 2021/09/01. doi: 10.1101/2021.08.15.21262077. PubMed PMID: 34462756; PubMed Central PMCID: PMCPMC8404894.

15. nCoV-2019 sequencing protocol [Internet]. (2020) [cited 28 JUL 2020].

16. Viral Amplicon Illumina Workflow (VAIW): A custom pipeline to analyze the SARS-CoV-2 genomes prepared with an amplicon (ARTIC (v3) and YouSeq (v2)) based library protocols [Internet]. (2020). Available from: https://hub.docker.com/r/bdrdgenomics/viral_amplicon_illumina_workflow.

17. Bushnell B. BBMAP: A Fast, Accurate, Splice-Aware Aligner. (2014). p. https://sourceforge.net/projects/bbmap.

18. Bushnell B, Rood J, Singer E. BBMerge -Accurate paired shotgun read merging via overlap. PLoS One (2017) 12(10):e0185056. Epub 2017/10/27. doi: 10.1371/journal.pone.0185056. PubMed PMID: 29073143; PubMed Central PMCID: PMCPMC5657622.

19. ARTIC Network Github [Internet]. (2020). Available from: https://github.com/artic-network/fieldbioinformatics/tree/master/artic.

20. Li H, Handsaker B, Wysoker A, Fennell T, Ruan J, Homer N, et al. The Sequence Alignment/Map format and SAMtools. Bioinformatics (2009) 25(16):2078–9. Epub 2009/06/10. doi: 10.1093/bioinformatics/btp352. PubMed PMID: 19505943; PubMed Central PMCID: PMCPMC2723002.

21. Grubaugh ND, Gangavarapu K, Quick J, Matteson NL, De Jesus JG, Main BJ, et al. An amplicon-based sequencing framework for accurately measuring intrahost virus diversity using PrimalSeq and iVar. Genome Biol (2019) 20(1):8. Epub 2019/01/10. doi: 10.1186/s13059-018-1618-7. PubMed PMID: 30621750; PubMed Central PMCID: PMCPMC6325816.

22. O’Toole A, McCrone, J.T., Scher E. Pangolin. GitHub (2020).

23. Katoh K, Misawa K, Kuma K, Miyata T. MAFFT: a novel method for rapid multiple sequence alignment based on fast Fourier transform. Nucleic Acids Res (2002) 30(14):3059–66. Epub 2002/07/24. doi: 10.1093/nar/gkf436. PubMed PMID: 12136088; PubMed Central PMCID: PMCPMC135756.

24. Minh BQ, Schmidt HA, Chernomor O, Schrempf D, Woodhams MD, von Haeseler A, et al. IQ-TREE 2: New Models and Efficient Methods for Phylogenetic Inference in the Genomic Era. Mol Biol Evol (2020) 37(5):1530–4. Epub 2020/02/06. doi: 10.1093/molbev/msaa015. PubMed PMID: 32011700; PubMed Central PMCID: PMCPMC7182206.

25. FigTree v1. 4 [Internet]. (2012). Available from: http://tree.bio.ed.ac.uk/software/figtree/.

26. Rambaut A, Holmes EC, O’Toole A, Hill V, McCrone JT, Ruis C, et al. A dynamic nomenclature proposal for SARS-CoV-2 lineages to assist genomic epidemiology. Nat Microbiol (2020). Epub 2020/07/17. doi: 10.1038/s41564-020-0770-5. PubMed PMID: 32669681.

27. Lubinski B, Frazier LE, Phan MVT, Bugembe DL, Tang T, Daniel S, et al. Spike protein cleavage-activation mediated by the SARS-CoV-2 P681R mutation: a case-study from its first appearance in variant of interest (VOI) A.23.1 identified in Uganda. bioRxiv (2021). Epub 2021/07/08. doi: 10.1101/2021.06.30.450632. PubMed PMID: 34230931; PubMed Central PMCID: PMCPMC8259907.

28. Thomas P. Peacock CMS, Jonathan C. Brown, Niluka Goonawardane, Jie Zhou, Max Whiteley, PHE Virology Consortium, Thushan I. de Silva, Wendy S. Barclay. The SARS-CoV-2 variants associated with infections in INdia, B.1.617, show enhanced spike cleavage by furin. bioRxiv (2021). doi: https://doi.org/10.1101/2021.05.28.446163.

29. Syed AM, Taha TY, Tabata T, Chen IP, Ciling A, Khalid MM, et al. Rapid assessment of SARS-CoV-2 evolved variants using virus-like particles. Science (2021):eabl6184. Epub 2021/11/05. doi: 10.1126/science.abl6184. PubMed PMID: 34735219.

30. B. Li Ad, K. Li, Y. Hu, Z. Li, Q. Xiong, Z. Liu, Q. Guo, L. Zou, H. Zhang, M. Zhang, F. Ouyang, J. Su et al. Viral Infection and Transmission in a large well-traced outbreak caused by the Delta SARS-CoV-2 variant. medRxiv (2021). doi: https://doi.org/10.1101/2021.07.07.21260122.

31. Ong SWX, Chiew CJ, Ang LW, Mak TM, Cui L, Toh M, et al. Clinical and virological features of SARS-CoV-2 variants of concern: a retrospective cohort study comparing B.1.1.7 (Alpha), B.1.315 (Beta), and B.1.617.2 (Delta). Clin Infect Dis (2021). Epub 2021/08/24. doi: 10.1093/cid/ciab721. PubMed PMID: 34423834; PubMed Central PMCID: PMCPMC8522361.

32. Teyssou E, Delagreverie H, Visseaux B, Lambert-Niclot S, Brichler S, Ferre V, et al. The Delta SARS-CoV-2 variant has a higher viral load than the Beta and the historical variants in nasopharyngeal samples from newly diagnosed COVID-19 patients. J Infect (2021) 83(4):e1–e3. Epub 2021/08/23. doi: 10.1016/j.jinf.2021.08.027. PubMed PMID: 34419559; PubMed Central PMCID: PMCPMC8375250.

33. Kissler SM, Fauver JR, Mack C, Olesen SW, Tai C, Shiue KY, et al. Viral dynamics of acute SARS-CoV-2 infection and applications to diagnostic and public health strategies. PLoS Biol (2021) 19(7):e3001333. Epub 2021/07/13. doi: 10.1371/journal.pbio.3001333. PubMed PMID: 34252080; PubMed Central PMCID: PMCPMC8297933 following competing interests: JW is an employee of Quest Diagnostics. JW is an employee of Bioreference Laboratories. NDG has a consulting agreement for Tempus and receives financial support from Tempus to develop SARS-CoV-2 diagnostic tests. SMK, SWO, and YHG have a consulting agreement with the NBA.

